# What does and does not correlate with COVID-19 death rates

**DOI:** 10.1101/2020.06.09.20126805

**Authors:** Christopher R. Knittel, Bora Ozaltun

## Abstract

We correlate county-level COVID-19 death rates with key variables using both linear regression and negative binomial mixed models, although we focus on linear regression models. We include four sets of variables: socio-economic variables, county-level health variables, modes of commuting, and climate and pollution patterns. Our analysis studies daily death rates from April 4, 2020 to May 27, 2020. We estimate correlation patterns both across states, as well as within states. For both models, we find higher shares of African American residents in the county are correlated with higher death rates. However, when we restrict ourselves to correlation patterns *within a given state*, the statistical significance of the correlation of death rates with the share of African Americans, while remaining positive, wanes. We find similar results for the share of elderly in the county. We find that higher amounts of commuting via public transportation, relative to telecommuting, is correlated with higher death rates. The correlation between driving into work, relative to telecommuting, and death rates is also positive across both models, but statistically significant only when we look across states and counties. We also find that a higher share of people not working, and thus not commuting either because they are elderly, children or unemployed, is correlated with higher death rates. Counties with higher home values, higher summer temperatures, and lower winter temperatures have higher death rates. Contrary to past work, we do not find a correlation between pollution and death rates. Also importantly, we do not find that death rates are correlated with obesity rates, ICU beds per capita, or poverty rates. Finally, our model that looks within states yields estimates of how a given state’s death rate compares to other states after controlling for the variables included in our model; this may be interpreted as a measure of how states are doing relative to others. We find that death rates in the Northeast are substantially higher compared to other states, even when we control for the four sets of variables above. Death rates are also statistically significantly higher in Michigan, Louisiana, Iowa, Indiana, and Colorado. California’s death rate is the lowest across all states.

*It is important to understand that this research, and other observational analyses like it, only identify correlations: these relationships are not necessarily* causal. *However, these correlations may help policy makers identify variables that may potentially be causally related to COVID-19 death rates and adopt appropriate policies after understanding the causal relationship*.

## 1 Executive Summary

We correlate county-level COVID-19 death rates (death per thousand people) with key variables using both mulitple linear regression and negative binomial mixed models, although we focus on linear regression models, the standard statistical tool used by economists. We include four sets of variables: county-level socio-economic variables, the modes of commuting, county-level health variables, and climate and pollution patterns. The socio-economic variables included are percentage of residents that are African American, Hispanic, white and other, the poverty share, the percentage of residents that are over 65, the percentage of people living in owner occupied housing, and the median home value. Our commuting mode variables include the share of workers driving, taking public transit, biking, walking, working from home, and not commuting at all (i.e., not working). Our health variables measure the share of smokers, diabetics, and uninsured in the county, obesity rates, and the number of ICU beds per capita. Finally, our climate and pollution variables include the average summer and winter temperatures and the level of air pollution, measured by the average amount of *PM*_2.5_ in the air.

It is important to understand that this study, and other observational analyses like it, only identify *correlations*: these relationships are not necessarily *causal*. That is, one cannot read this study, or others like it, to imply that changing one of the variables in our model would change death rates; we can only say how the death rates and the variables analyzed move together. We take much care in stressing this throughout the note. While causal relationships are often needed to base policy upon, the correlations we identify may help policy makers identify variables that may potentially be causally related to COVID-19 death rates and adopt appropriate policies after understanding the causal relationship. Additionally, it is important to note the potential gap between true COVID-19 deaths and the deaths data that is available and consequently used in this study. This gap, which is a result of a county’s ability to capture or miss deaths, quite likely varies between each county as well, depending on different testing capacities. As a result, some of the correlations we uncover may reflect correlations between the variables in our model and reporting errors.

The multiple regression framework used in our analysis does, however, allow researchers and policy makers to better focus their attention on potential causal mechanisms that might be driving the correlations we uncover. The way to interpret the correlations we estimate is that these are the correlations between death rates and a given variable, say the share of residents that are African American, *after controlling for the other variables in the model*. So, for example, if we find a positive relationship between African Americans and death rates, we can say that this relationship is *not* driven by income and/or health insurance disparities because we control for income and health insurance in the model. The causal mechanism has to be some factor that is not already in our statistical model. Therefore, our framework can not only say what is and what is not correlated with death rates, but also allows policy makers to eliminate potential channels driving these important correlations.

Our analysis studies county-level daily death rates from April 4, 2020 to May 27, 2020. We estimate separate statistical models for each day and show the patterns of correlations over time. We note that we omit the five counties that comprise New York City in our main analysis. We include the results that include these counties in the Appendix.

We estimate linear regression models both with and without what are known as “state fixed effects.” State fixed effects are a set of indicator (dummy) variables, one for each state, that allow each state to have their own baseline death rates. These differences in baseline death rates capture variables that are common across counties within the same state. Which state we choose as the comparison state is arbitrary and does not affect the relative death rates across any two states. We choose Alabama. Because these measure the relative death rates across states after controlling for their socio-economic variables, health variables, commute patterns, and climate and pollution patterns, one can interpret these as whether a given state is fairing better or worse in terms of death rates, conditional on these variables.

The inclusion of state fixed effects also changes the interpretation of the other variables included in the model. When state fixed effects are not included, the correlations are measured using variation in death rates and the variables included in the model both within a state and *across* states. However, when the state fixed effects are included, only variation across counties *but within the same state*, is used to measure the correlation. The correlation patterns across these two models are important because often one might find two variables correlate with each other across states, but not within. In cases like these one should worry that the former correlation is driven by *other* factors, common to a given state, not accounted for in the statistical model.

In the models that do not control for the state fixed effects, A higher share of African Americans rates are correlated with higher death rates; this correlation remains positive when we control for the state, but is no longer statistically significant. To give a sense of the magnitude of the correlation, the coefficient (without state effects) at the end of the sample implies that deaths per 1,000 people are, on average, 1.262 higher in a county that has all African American residents compared to a county that has no African American residents. The average county-level death rate at the end of the sample is 0.119 (omitting New York City). One useful thought experiment is to consider the death rates in two “average” counties one with an African American share of the lowest county in our data (0%) and one with an African American share equal to the largest we observe (87%). Our results suggest that moving between these two counties would be associated with an increase in the death rate of 1.10 per 1,000 residents, nearly a 10-fold increase. The coefficient with fixed state effects implies more than a tripling in the death rate; however, we stress that this is not statistically significant. Given, the importance of this issue, we encourage policy makers and researchers to investigate the causal relationship between death rates and the share of African American residents.

As noted above, this correlation is not driven by any combination of the other variables in our model. For example, a natural instinct among policy makers may be to think this correlation is because of income disparities. It is not. We control for income in our multiple regression model, so any income disparities between African Americans and other races would be *additional* to the correlation we uncover. Similarly, we control for health insurance status, diabetes, poverty rates, obesity, smoking rates, and public transit use. Therefore, the correlation we uncover between death rates and the share of African Americans *cannot* be driven by these other variables. That is to say, the reason why African Americans face higher death rates is not because they have higher rates of uninsured, poverty, diabetes, etc.; it must be some other mechanism. For example, it could be because the quality of their insurance is lower, the quality of their hospitals is lower, or some other systemic reason. Our analysis can hopefully allow policy makers to focus on a narrower set of potential causal links.

We find higher rates of diabetes in the county is correlated with higher death rates whether we control for the state or not; however, this relationship is no longer statistically significance when controlling for state, but remains positive. A higher share of elderly in the county are correlated with higher death rates whether we control for the state or not, although the statistical significance is marginal. Finally, we find that counties with higher home values, higher summer temperatures, and lower winter temperatures have higher death rates.

A striking and robust relationship is found between death rates and public transit use. We find higher rates of commuting via public transportation is associated with higher death rates compared to all other modes of commuting, including not working, whether we control for state fixed effects or not. This correlation is statistically significant when comparing with telecommuting. Counties with a higher share of workers driving and walking, relative to telecommuting, also have statistically significantly higher death rates. Taken together, these results suggest that counties with high levels of telecommuters have lower death rates.

The magnitude of the public transportation correlation is large. The share of people commuting via public transportation ranges from 0 to 20.6 percent across the counties in our sample (again, omitting New York City counties). The coefficient from the model with fixed state effects implies that a 20.6 percentage point increase in public transportation use (relative to telecommuting), the range in public transit use in our data, is associated with a 0.99 increase in the death rate per 1,000 people. This *increase* is nearly 10 times the average death rate across all counties of 0.119.

One can speculate as to what might be driving these correlations. We uncover two stylized facts. First, the mode of commute correlated with the highest death rates is public transit. Second, depending on which model we focus on, there is evidence that all modes of commutes, other than biking, are associated with higher death rates relative to telecommuting. A scenario that is consistent with both of these stylized facts is that some of higher death rate associated with public transit use is coming from public transit itself. And, some is coming from the day-to-day interaction with others as part of the types of jobs those workers taking public transit do. This second effect is also present for those walking and driving to work and the reason they face higher rates; these workers also have more interaction with others compared to telecommuting. Combined, if this speculation is correct, it may point to a greater need to disinfect and socially distance public systems and a greater need for social distancing at work.

This story does not explain the positive correlation between death rates and the “other” mode share, which represents those not working—elderly, children, and the unemployed.^1^ It is not likely the higher death rates associated with other is because of elderly given that we control for the share of elderly in the model. Therefore, this positive correlation is likely the result of a greater number of children and unemployed. Perhaps, for example, the causal link is do to those counties with greater children have larger social networks and, thus, higher exposure rates, and the stress of unemployment is an additional causal mechanism.

Also importantly, we do *not* find that death rates are correlated with local pollution, obesity rates, ICU beds per capita, or poverty rates. The lack of a correlation with pollution levels contrasts with recent work by Wu et al. (2020). We explore what is driving this difference in section 4. This analysis suggests that once additional health and commute mode variables are included, the size of the pollution correlation falls and statistical significance goes away, suggesting that the correlation between death rates and pollution may be spurious.

Our model that looks within states yields estimates of how a given state’s death rate compares to other states after controlling for the variables in our model; this may be interpreted as a measure of how states are doing relative to other states. We find that death rates in the Northeast are substantially higher even when we control for the four sets of variables above. Death rates are also statistically significantly higher in Michigan, Louisiana, Iowa, Indiana, and Colorado. Accounting for the variables in our model, California has the lowest death rate compared to all other states.

## 2 Data and Methods

We discuss the data sources and methodology here. The COVID-19 deaths are collected from the COVID-19 Data Repository by the Center for Systems Science and Engineering (CSSE) at Johns Hopkins University. County-level data and the source of the data can be found in Table 1^2^.

**Table 1:** Coefficients for mean *PM*_2.5_ with Deaths as dependent variable

| Dataset | Source |
| --- | --- |
| Obesity and Diabetes | 2016 Diabetes Surveillance system |
| Smoking rate | Average of 2016, 2015, 2014, and 2006-2012 Behavioral Risk Factor Surveillance System |
| Uninsured | 2017 Small area health insurance estimates |
| Transportation | American Community Survey Data (5-year estimate using data from 2015) |
| $PM_{2.5}$ | The Atmospheric Composition Analysis Group at Dalhouse University (via Wu et al. (2020)) |
| Percent ICU Beds | Kaiser Health News |
| Poverty | American Community Survey Data (via Wu et al. (2020)) |
| Owner Occupation | American Community Survey Data (via Wu et al. (2020)) |
| Demographics | American Community Survey Data (via Wu et al. (2020)) |
| Mean temperature | Gridmet via Google Earth Engine (via Wu et al. (2020)) |
| Population | American Community Survey Data (via Wu et al. (2020)) |
| Population density | American Community Survey Data (via Wu et al. (2020)) |
| Education | American Community Survey Data (via Wu et al. (2020)) |
| Income | American Community Survey Data (via Wu et al. (2020)) |
| COVID-19 Deaths | Johns Hopkins University |

Our county-level variables fall into four general categories:

- **Socio-economic Variables:** Percent of population below the poverty line, percent of residents that are African American, percent white, percent Hispanic, percent old, percent owner occupied housing, *log*(median house value), dummy variables for five income bins, dummy variables for four education levels, dummy variables for four population density bins.
- **Commute patterns:** Share of people that drive, use public transit, bike, walk, and work from home
- **Health:** Percent smokers, obese, diabetic, ICU beds, uninsured
- **Climate and Pollution:** *log*(mean summer temperature), *log*(mean winter temperature), mean *PM*_2.5_

The sources for the data are reported in Table 1. We have data for 3,033 counties. For our main analysis, we omit the five counties that comprise New York City, which leads to a total of 3028 counties after taking out New York City. For those that are interested, replicates of the Figures in the main section that include New York City can be found in the Appendix section.^3^ The data have cumulative deaths at each given day between April 4th and May 27th.

## 3 Results

### 3.1 Transportation, health, socio-economic, climate and pollution

We present the results in table and figure format. Figures 3 through 6 in Section 5 plot the coefficients of interest across the four sets of variables in the regression models. The left panel plots the coefficients for the model without fixed state effects, while the right panel plots the coefficients when we include fixed state effects. We estimate the regression model each day; the dark blue line representing the coefficient estimate and blue shaded area the 95-percent confidence interval. Table 2 in Section 6 reports the regression results for three specific days, April 4, April 30, and May 27.

Prior to getting to the specific results, we again stress that these are correlations; they do not necessarily imply that the relationships are causal. That is, our analysis and those like ours that focus on observational data, can understand how death rates move with certain variables, but that movement does not imply that the variables we analyze *cause* higher or lower death rates. Other variables, not in the analysis, may be the causal link. However, we do feel that the correlations that we uncover may help policy makers know where to focus when trying to determine causal relationships. Furthermore, the correlations themselves may be important pieces of data for policy makers.

**Table 2:**
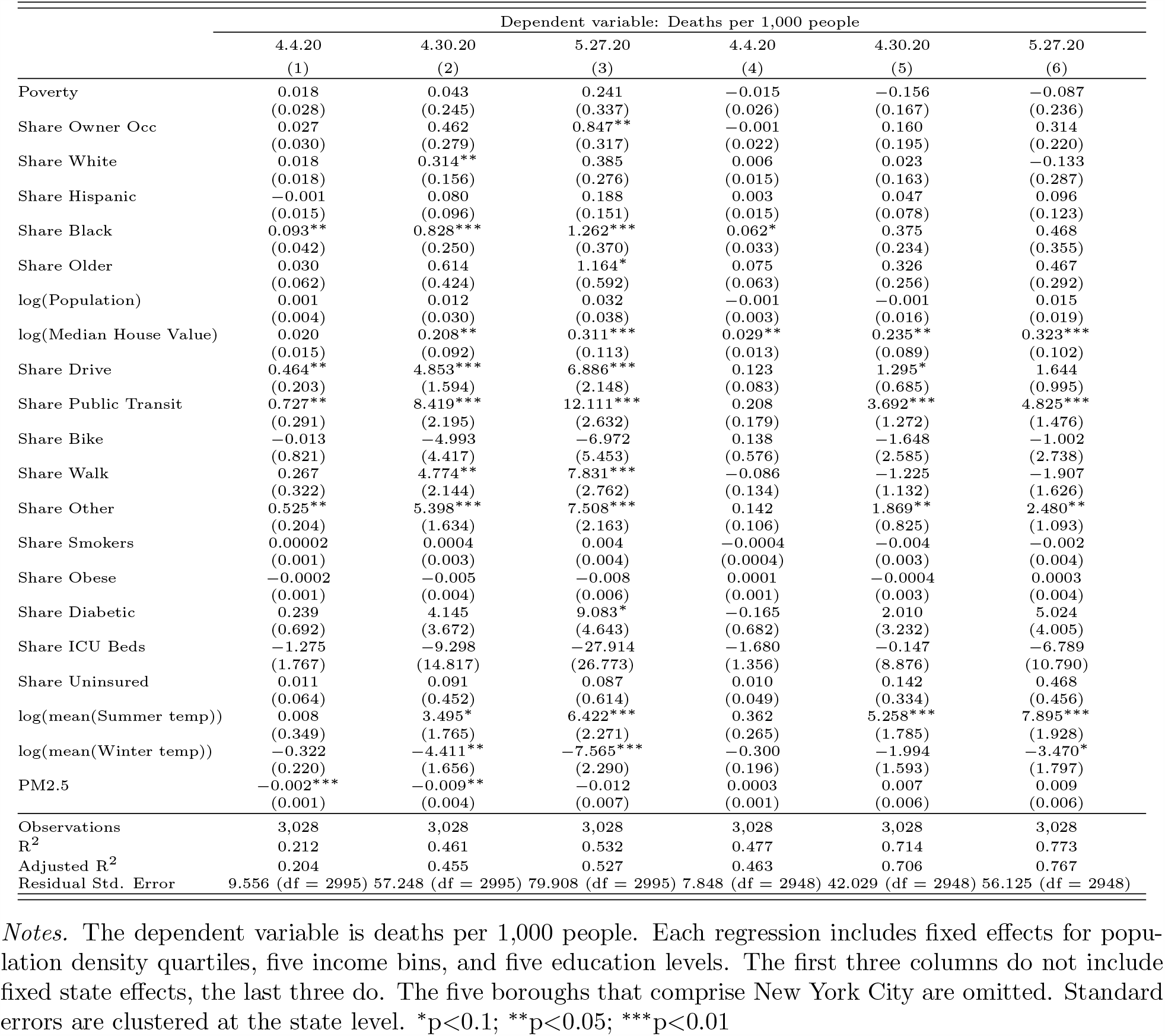
Main Regression table, first three columns are without states last three are with states

We begin by discussing three correlations that have received significant attention: the relationship between COVID19 deaths and the share of the county’s population that is African American residents, public transit reliance, and pollution.

We find a positive, statistically significant, and large correlation between death rates and the share of residents that are African American in the model without state fixed effects, but the correlation is no longer statistically significant when we include fixed state effects. To give a sense of the magnitude of the correlation, the coefficient (without state effects) at the end of the sample implies that a 10 percentage point increase in the share of residents that are African American is associated with an increase of 0.126 (0.1·1.262) in deaths per 1,000 people. Considering that the average death rate at the end of our sample is 0.119, this is a more than doubling in the average death rate. Another useful thought experiment is to use the range in the share of African Americans that we observe in the data. Within our data, the African American share ranges from zero to 87%. Consider the death rates in two “average” counties one with an African American share of the lowest county in our data and one with an African American share equal to the largest we observe. Our results suggest that moving between these two counties would be associated with an increase in the death rate of 1.10 per 1,000 residents (0.87·1.262), nearly a 10-fold increase.

It is important to note, however, that the statistical significant of this correlation goes away when we control for fixed state effects. The coefficient with fixed state effects remains large, implying more than a tripling in the death rate, but the lack of statistical precision means we cannot rule out there is no relationship. This makes the relationship between death rates and the share of African American residents more nuanced. In particular, this suggests that the correlation in the model that does not account for state effects is driven by differences in death rates and the share of African Americans *across* states and not within states. That is, once we account for the state within the statistical model, the share of African American residents is no longer associated with death rates. For example, our results are consistent with the scenario where states with a higher share of African Americans have higher death rates than those states with a lower share, say Louisiana versus Tennessee, but those counties within Louisiana that have a higher share of African Americans do not have higher death rates compared to those counties within Louisiana that have a lower share of African Americans. These conflicting results underscore the importance of fully understanding the causal relationship between race and death rates.

We next turn to the use of public transit. Higher public transit use is correlated with higher death rates in both the models without fixed state effects and the models with fixed state effects. The omitted commute mode is telecommuting, so the correlation between public transit use and the death rate should be interpreted as relative to the scenario where those taking public transit instead work from home. We also remind the reader that we have omitted the five counties that comprise New York City from the analysis. Including them makes this correlation much stronger.

The magnitude of these correlations is very large. At the end of our sample, the coefficients associated with this variable are 12.114 and 4.822 in the models without and with fixed state effects, respectively. Therefore, a 10 percentage point increase in public transit use, relative to telecommuting, is associated with a 1.21 (0.1·12.11) and 0.48 (0.1·4.82) increase in deaths per 1000 people in the model without and with state fixed effects, respectively. Public transit share ranges from zero to 20.6% across counties. Using the same thought experiment as above, consider two “average” counties one with a public transit share of zero and one with a public transit share of 20.6%. Our results suggest that moving between these two counties would be associated with an increase in the death rate of 2.49 per 1,000 residents (0.206·12.11) and 0.99 per 1,000 residents (0.206·4.83), respectively; an increase of over 21-fold and 8-fold, respectively.^4^

We note that there is also a positive relationship between driving to work, walking to work, and not commuting at all (“other”) and deaths per 1,000 people when we look across states, but the correlation is no longer statistically significant for walking and driving when we look within state. Taken together these results suggest that telecommuting is correlated with lower death rates, compared to most other modes of commuting and that public transit use is associated with the highest death rates.^5^ We discuss this further below.

The role of pollution in COVID-19 deaths has received significant attention in the popular press.^6^ We do not find a statistically significant effect of pollution on death rates. Furthermore, in the model without fixed state effects, we find a negative (but statistically insignificant) relationship. Given the analysis of Wu et al. (2020), we explore what is driving the differences in the results in Section 4.

Our results confirm a number of correlations that have been noted by public health officials. In the model without fixed state effects, by the end of the sample there is a statistically significant positive correlation between the share of diabetics in the county and death rates. Similar to the coefficients associated with the share of African Americans in the county, when we control for the state, this relationship is no longer statistically significant. The share of the population that is above 65 years of age is also positively associated with death rates; this association is marginally statistically significant (at the 10% level) in the model without state effects and not significant in the model with state effects.

Interestingly, the median house value is positive associated with death rates. We remind the reader that we are also controlling for average income in the county, so one should interpret the positive correlation with home values as: when comparing two counties with similar incomes, but different home values, death rates are higher in the county with higher home values. We also find statistically significant correlations between a county’s climate and death rates. In particular, higher average summer temperatures are associated with higher death rates, but higher average winter temperatures are correlated with lower death rates. This holds even within a state, so the correlation is not capturing, for example, the tragic loss of life in the Northeast or any other specific region.

We do not find clear patterns with any of the additional variables included in the analysis. These include: obesity, uninsured, poverty rates and the number of ICU beds per capita.

### 3.2 Summary of correlations

Figure 1 summarizes the strength of the correlations between death rates and the variables included in the model. We follow the calculations discussed above for each variable. Specifically, we take the range of each variable observed in our data across counties and multiply that range by the coefficient associated with that variable. The left bar represents this calculation for the model without state effects, while the right bar represents the calculation for the model with fixed state effects. The standard errors of the calculations are also shown. We order the variables by the magnitude of the calculation in the model without state effects. Viewing the coefficients in this way illustrates the strength of the correlation with public transit. The magnitude of the association with the share of diabetics is also large. The share of people not commuting to work, which likely represents retirees and children, and the share driving to work are also strongly correlated in both models.

**Figure 1:**
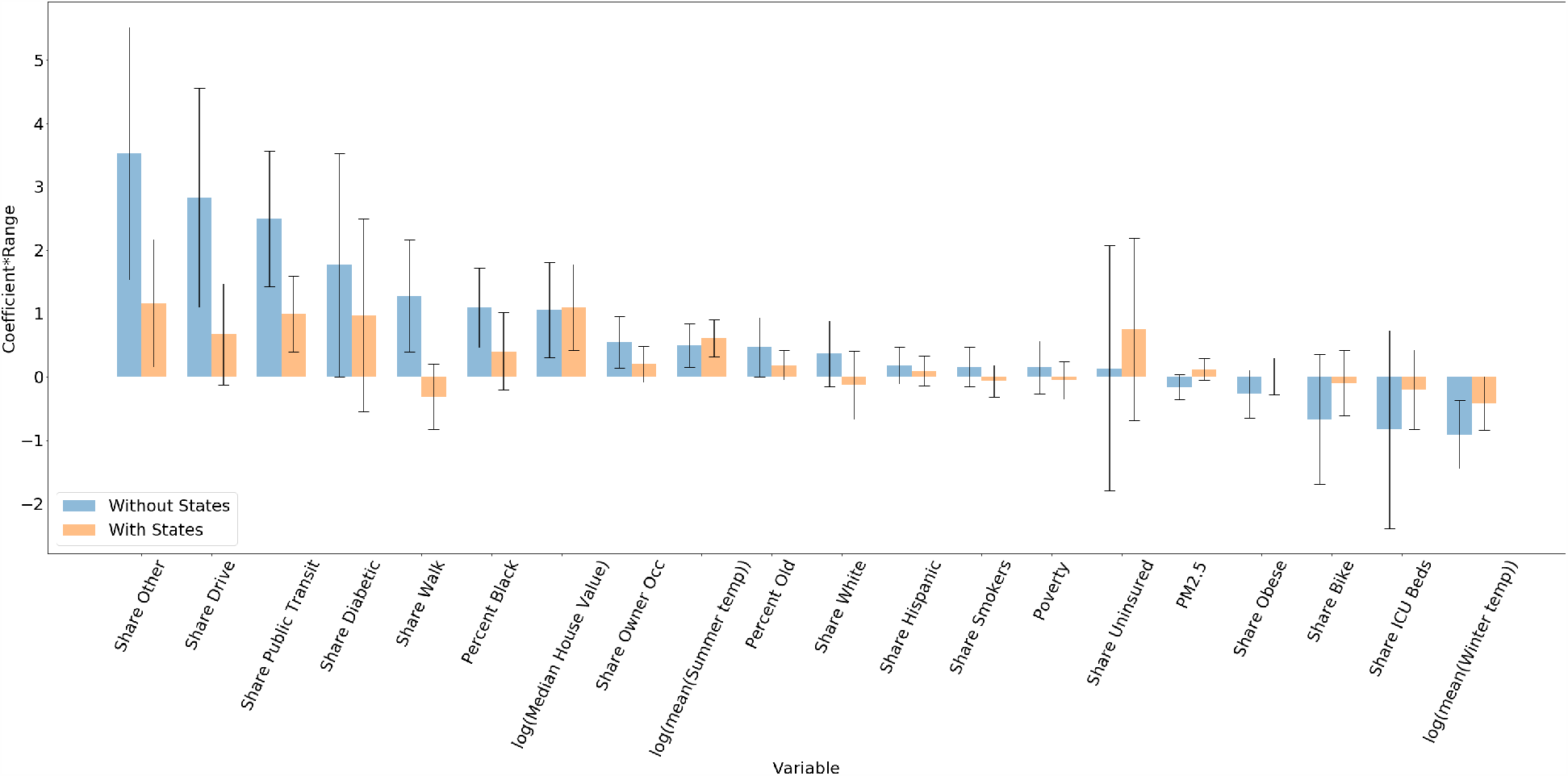
Bar graph of the estimate coefficient multiplied by the observed range of the associated variable across counties with 95 % CI.

Combined the mode of commute correlation pattern may help policy narrow in on potential causal mechanisms. Our results are consistent with two stylized facts. First, for a given percentage point change in its value, the mode of commute correlated with the highest death rates is public transit. This comes directly from Table 2. Second, depending on which model we focus on, there is evidence that all modes of commutes, other than biking, are associated with higher death rates relative to telecommuting. While this is speculative for the reasons we discussed above, one scenario that is consistent with both of these stylized facts is that some of higher death rate associated with public transit use is the result of public transit use itself. And, some is the result of what these workers are exposed to after they get to work. This second effect explains why those walking and driving to work also face higher rates; these workers also have more interaction with others compared to telecommuting. Combined, *if* this speculation is correct, it may point to a greater need to disinfect and socially distance public systems and a greater need for social distancing at work.

What, then, is the reason for higher death rates among counties with higher shares of “other” mode share, which represents those not working—elderly, children, and the unemployed? We control for the share of elderly in the county, so it is unlikely driven by this. Therefore, this positive correlation is likely the result of a greater number of children and unemployed. Perhaps, for example, the causal link is do to those counties with greater children have larger social networks and, thus, higher exposure rates, and the stress of unemployment is an additional causal mechanism.

The correlations with weather conditions are interesting and may warrant further investigation to understand their causal relationships with the virus. Higher summer temperatures and lower winter temperatures are correlated with higher death rates. This is true even when we look within a state.

We again point to the importance of understanding the causal relationship between death rates and the share of African Americans living in the county. While the magnitudes of this calculation are not as large as some of the other variables, and the correlation is statistically insignificant in the model with state fixed effects, the magnitude of the correlation in the model without state effects, implies a county with the highest observe share of African Americans is associated with an almost 10-fold increase in their death rate, relative to a county with the lowest share of African Americans. Understanding what is driving this correlation is an important public policy issue.

As noted above, this correlation is not driven by any combination of the other variables in our model. For example, a natural instinct among policy makers may be to think this correlation is because of income disparities. It is not. We control for income in our multiple regression model, so any income disparities between African Americans and other races would be *additional* to the correlation we uncover. Similarly, we control for health insurance status, diabetes, poverty rates, obesity, smoking rates, and public transit use. Therefore, the correlation we uncover between death rates and the share of African Americans *cannot* be driven by these other variables. That is to say, the reason why African Americans face higher death rates is not because they have higher rates of uninsured, poverty, diabetes, etc. it must be some other mechanism. For example, it could be because the quality of their insurance is lower, the quality of their hospitals is lower, or some other systemic reason. Our analysis can hopefully allow policy makers to focus on a narrower set of potential causal links.

### 3.3 Relative death rates across states

As noted above, the estimated coefficients on each state indicator variable measures how the average death rate in counties within a particular state compares to the average death rate across counties in Alabama, after controlling for the variables included in the model (e.g., the four sets of variables included). While the coefficients directly measure how a state compares to Alabama, one can take the difference between any two coefficients to compare any two states. These coefficients are plotted for the last day of our sample in Figure 2 along with confidence intervals. Perhaps not surprisingly, our results suggest that death rates are higher in the Northeast relative to other parts of the country even after controlling for the variables in our model. In addition to the Northeast, we find higher death rates in Michigan, Louisiana, Iowa, Indiana, and Colorado as well.

**Figure 2:**
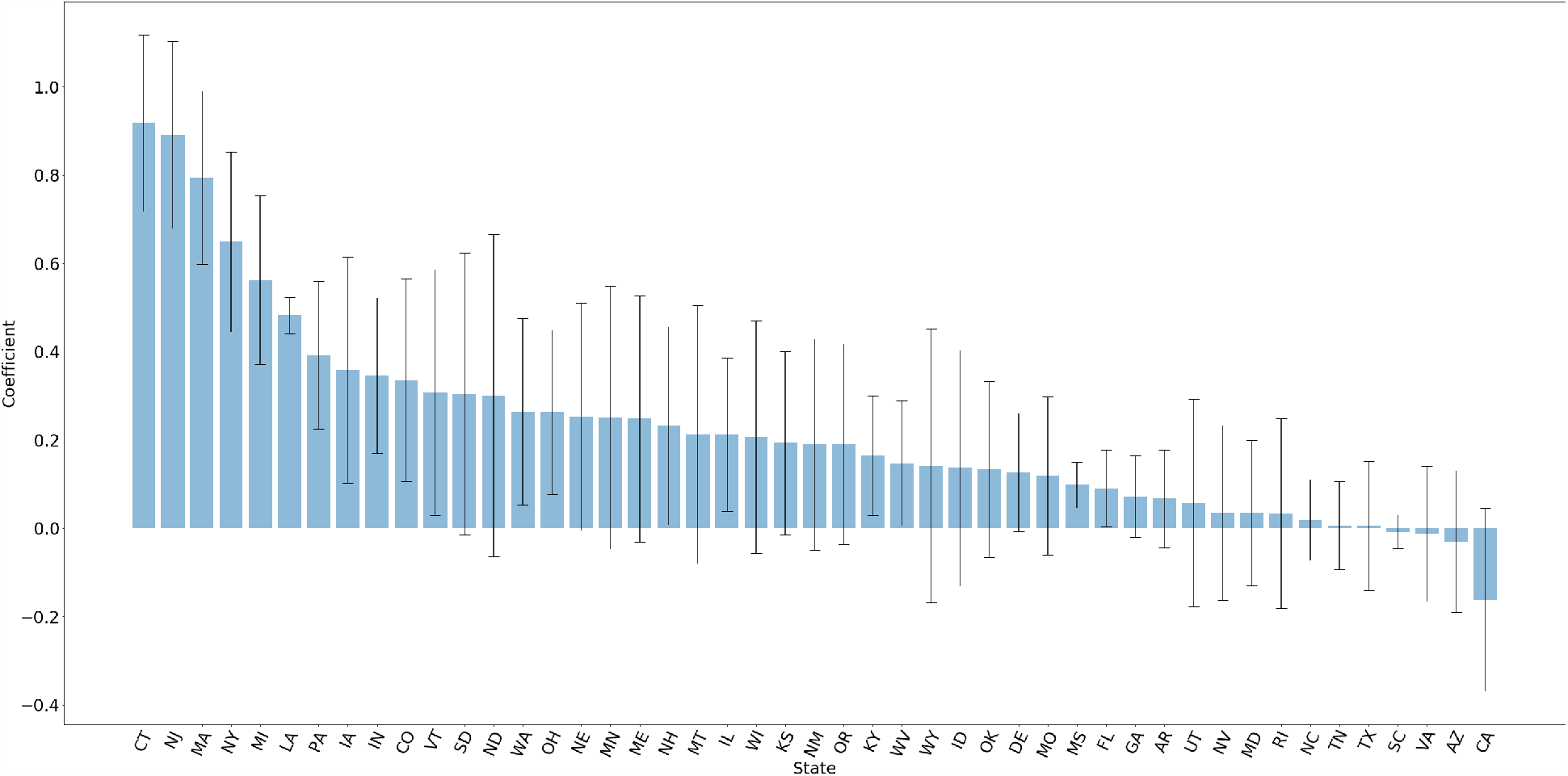
Bar graph of state fixed effects for the last day in the dataset with 95 % CI.

**Figure 3:**
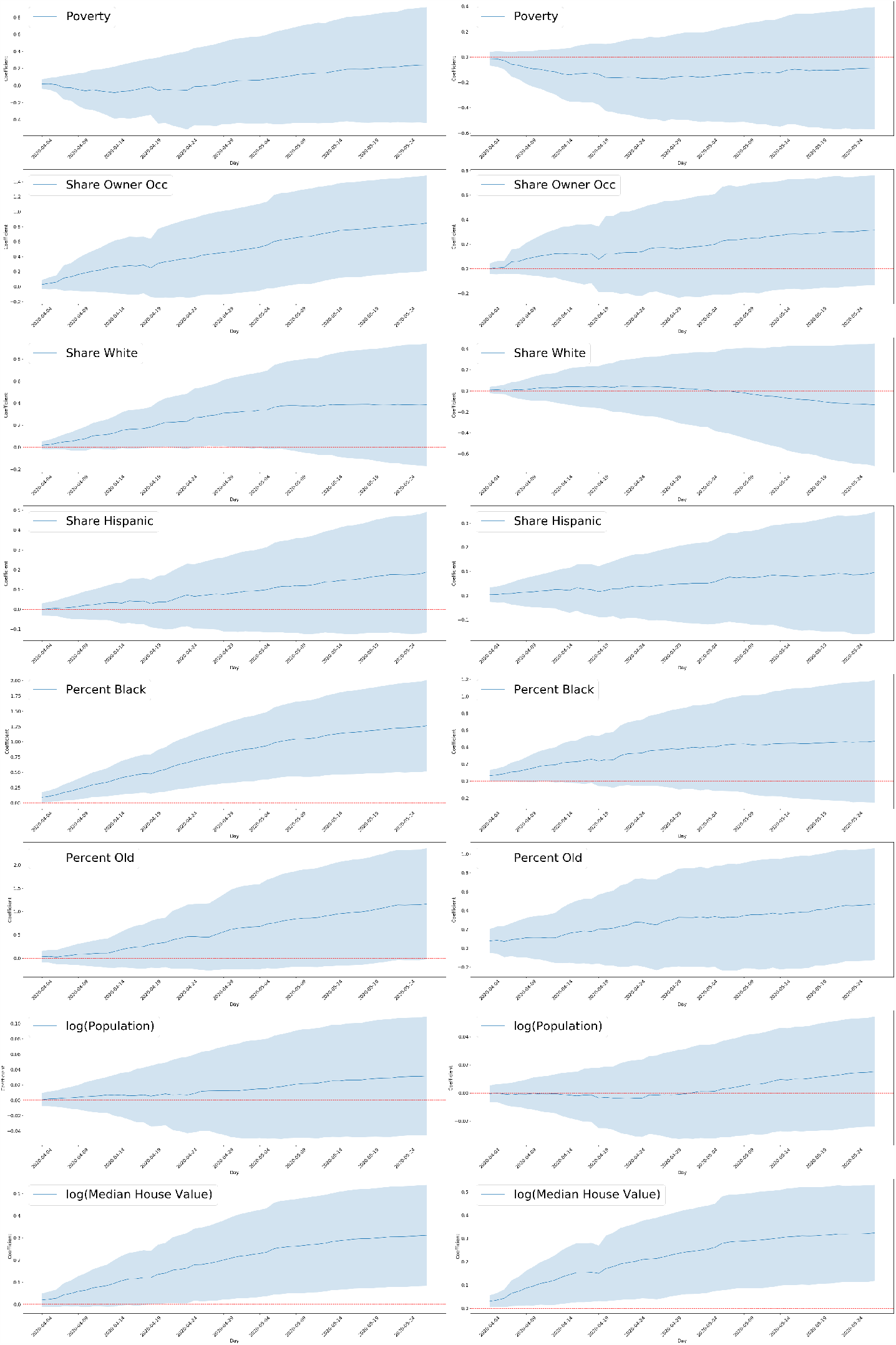
Each row represents a variable from the socio-economic category with the 95% CI associated with that variable. The first column is for the regressions run without state fixed effects and the second column is for the regressions run with state fixed effects. The x-axis represents the day and the associated coefficient is the regression run on the cumulative deaths on that given day. The horizontal red line represents 0.

**Figure 4:**
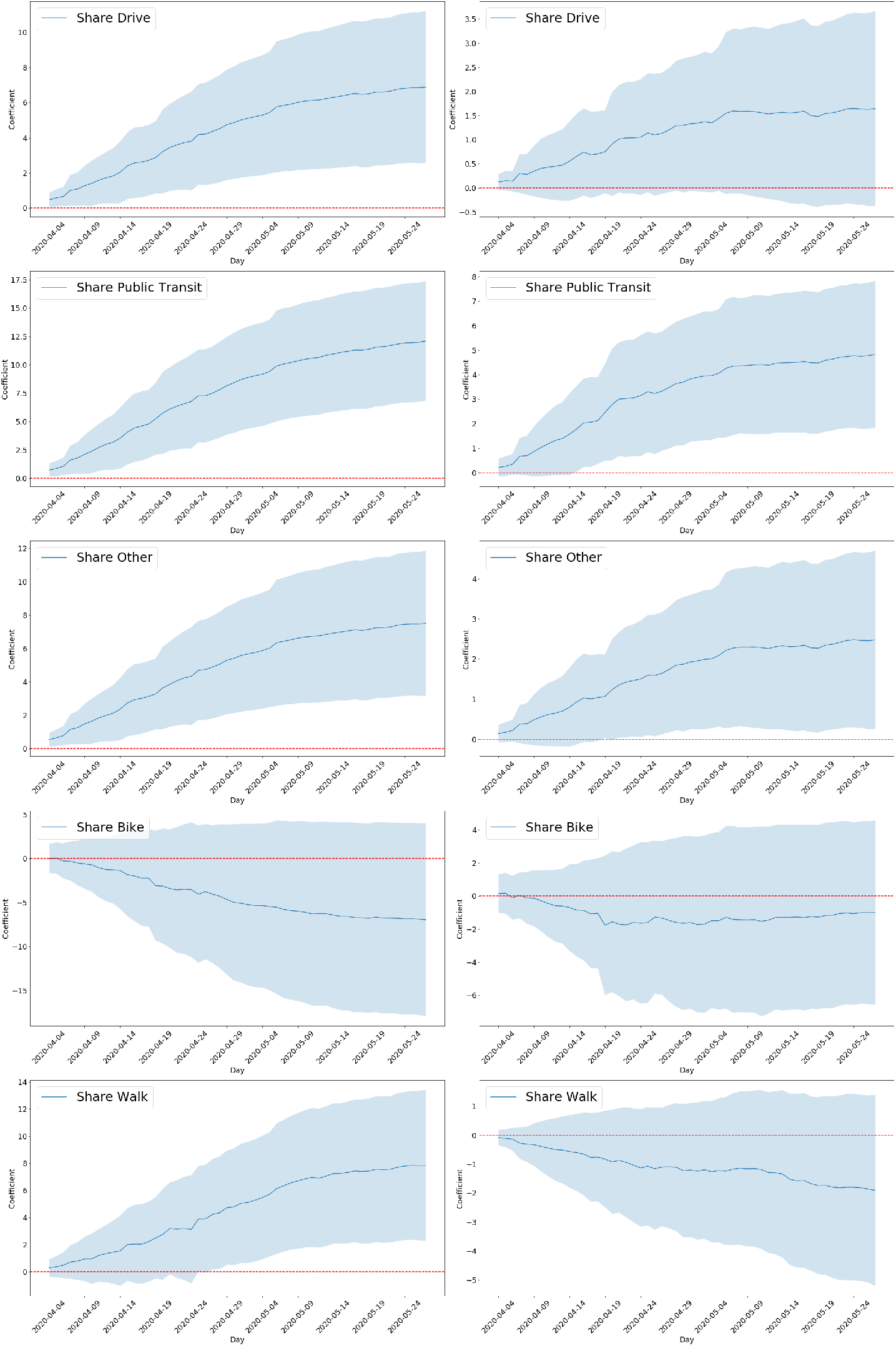
Each row represents a variable from the commute patterns category with the 95% CI associated with that variable. The first column is for the regressions run without state fixed effects and the second column is for the regressions run with state fixed effects. The x-axis represents the day and the associated coefficient is the regression run on the cumulative deaths on that given day. The horizontal red line represents 0.

**Figure 5:**
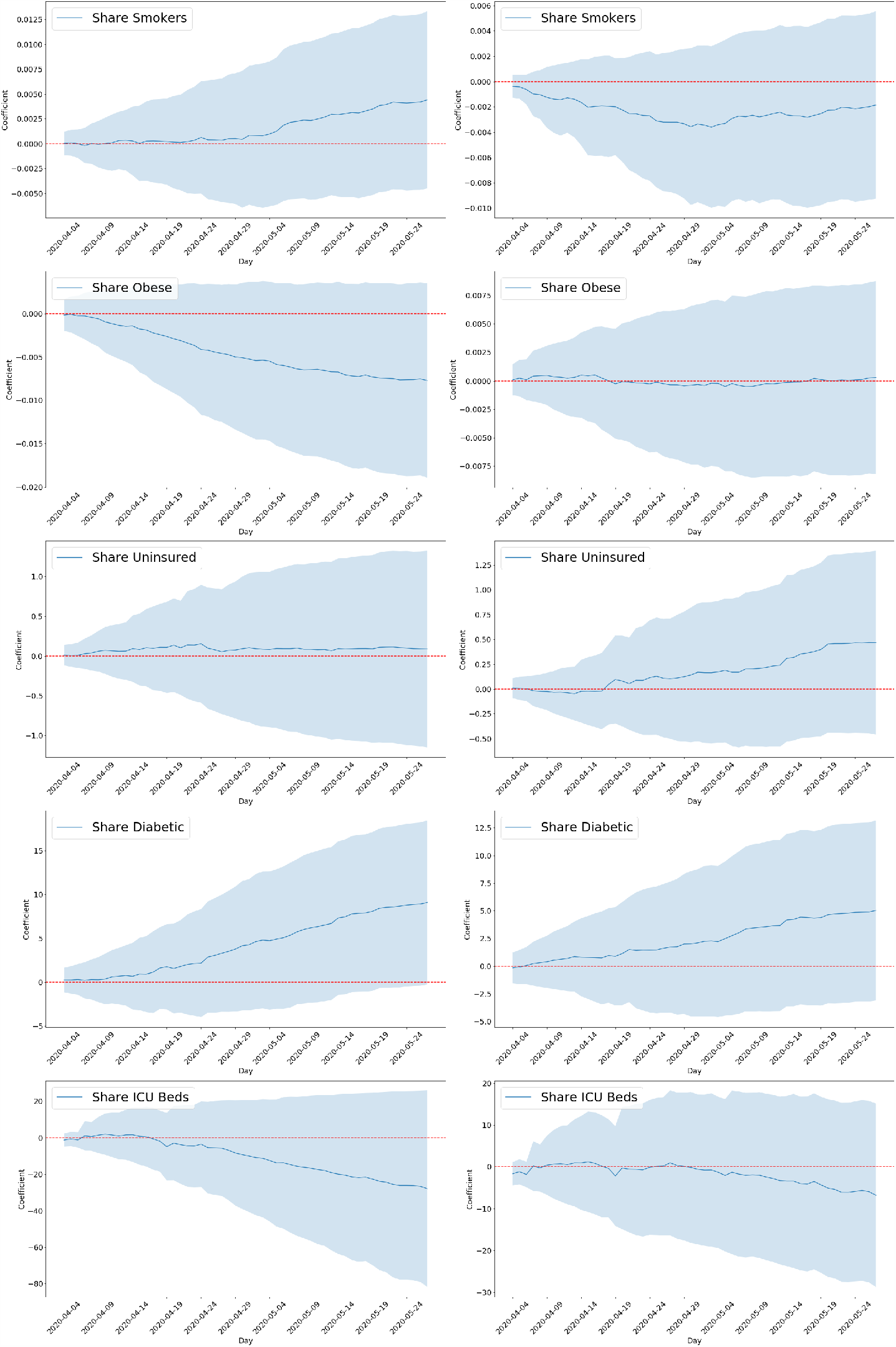
Each row represents a variable from the health category with the 95% CI associated with that variable. The first column is for the regressions run without state fixed effects and the second column is for the regressions run with state fixed effects. The x-axis represents the day and the associated coefficient is the regression run on the cumulative deaths on that given day. The horizontal red line represents 0.

**Figure 6:**
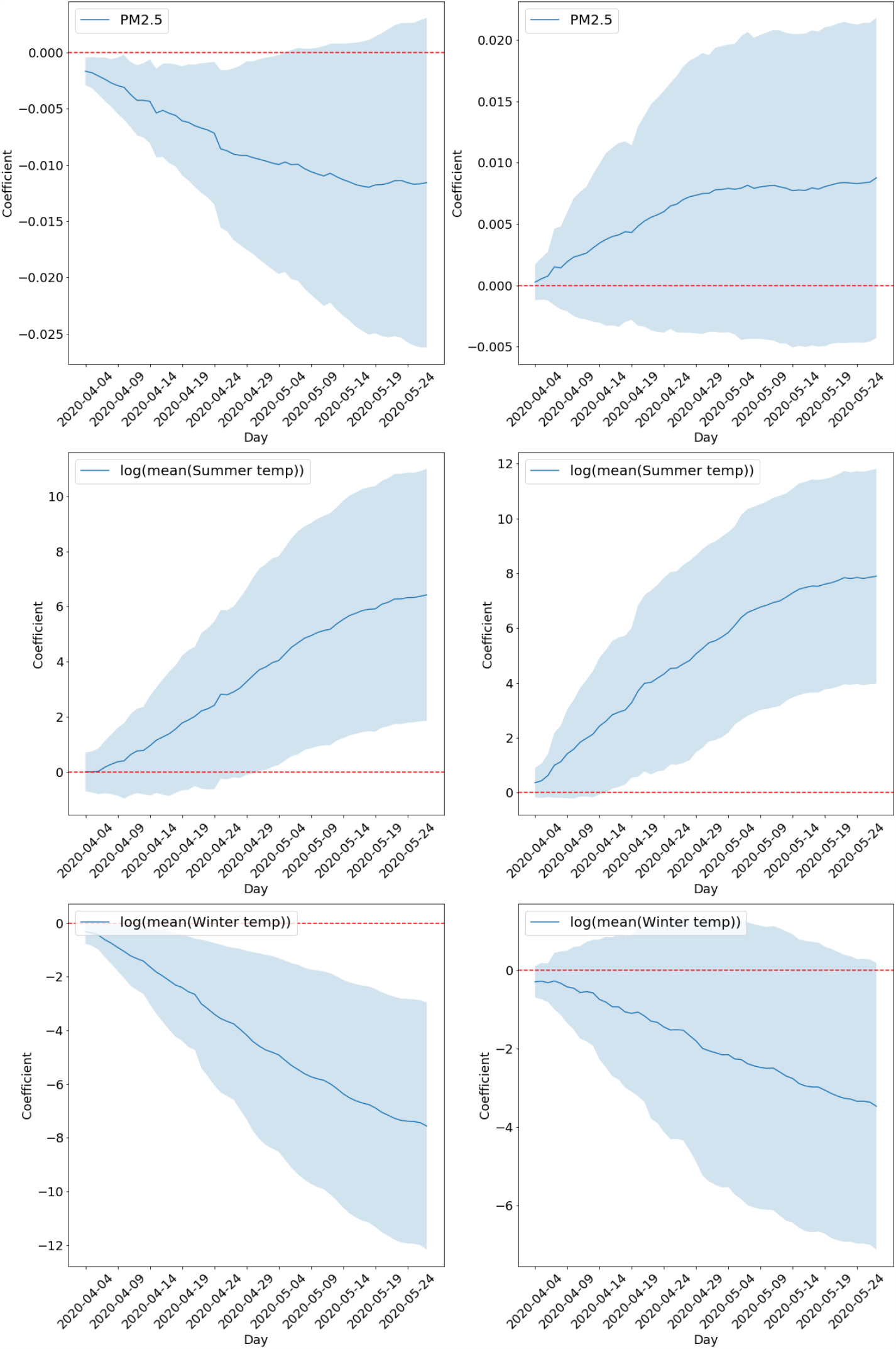
Each row represents a variable from the climate and pollutant category with the 95% CI associated with that variable. The first column is for the regressions run without state fixed effects and the second column is for the regressions run with state fixed effects. The x-axis represents the day and the associated coefficient is the regression run on the cumulative deaths on that given day. The horizontal red line represents 0.

The magnitude of these coefficients is large. For example, the Connecticut coefficient suggests that controlling for the variables included in our model, death rates in Connecticut are 0.9 higher than death rates in Alabama. Given an average county-level death rate of 0.12, this is a significant difference.

Across all states, California has the lowest death rate, after accounting for the variables included in the regression model. The coefficient implies deaths per 1000 are 0.16 lower in California, compared to in Alabama, once you control for the variables in the model. Figure 7 plots the state fixed effect coefficients over time. They are extremely stable in terms of their relative values. They will naturally increase in magnitude because death rates are increasing over this time period.

**Figure 7:**
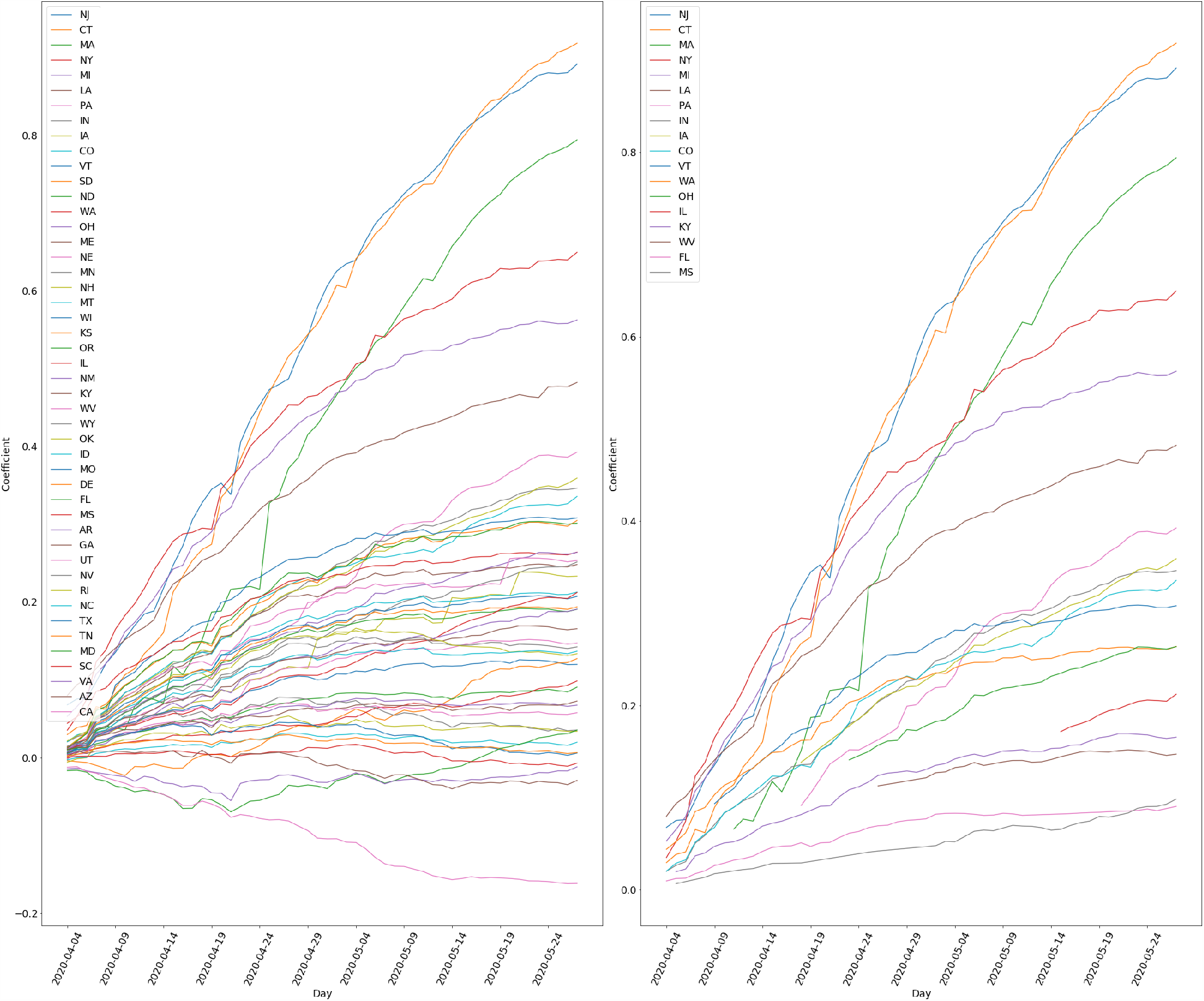
This figure plots the coefficient associated with each state fixed effect over time. The subfigure plots the coefficients for all states, the subfigure to the right plots only those that are statistically different from the omitted state, AL.

## 4 Comparison of our results with Wu et al. (2020)

Wu et al. (2020) has had two main versions to our knowledge, which will be referred to as the April 4 and April 24 versions. From these versions, we will only look at their main model, but note that they have run many variations of their model in both versions. Our analysis differs from Wu et al. (2020) in a number of ways. First, the number of counties differs from both their versions. The April 4 version’s main analysis restricts the model to 1,785 counties, their updated analysis runs on 3,089 counties.^7^ Our final analysis uses 3,028 counties.^8^ Second, Wu et al. (2020) focuses on the number of deaths, while we focus on deaths per 1,000 people. Because of their focus on deaths and the presence of a number of low numbers in some counties, Wu et al. (2020) estimate a Negative Binomial Mixed Model (NBMM). Fourth, the variable differ between the two sets of analyses.^9^ Fifth, the analysis of Wu et al. (2020) ends on May 5, 2020^10^, while our’s continues to May 27, 2020. Finally, within the regression models we estimate, we weight counties by their population. Within our data the county-level population varies from 63 to over a million (ignoring NYC). Not weighting the county-level observations by population treats all counties equally.

Table 3 is our attempt to understand what is driving the differences in the results with respect to the *PM*_2.5_ variable. In particular, we vary the differences with Wu et al. (2020) on a one-on one-off basis to do this. We report the estimate and the 95% confidence interval of that estimate for the last day of our analysis. The first row in the table reproduces the results in the April 4 version of Wu et al. (2020). The next row reproduces the results in the April 24 version of Wu et al. (2020), which adds more counties to the analysis by changing the covariates used in the analysis. The updated version’s estimate is lower, but still significant. In the third row, we use our baseline control variables.^11^ Here, the size of the coefficient falls significantly compared to the April 4 version and is similar to the April 24 version, but somewhat smaller and not statistically significant. Next, we add both our additional health and transport variables. The coefficient becomes even smaller and remains statistically insignificant (i.e., zero is not contained in the confidence interval). Combined, these results suggest that when we continue to define the dependent variable as deaths and estimate the same statistical model as in Wu et al. (2020), the statistically significant relationship between pollution and deaths no longer exists when we move to our setup which in its final form adds additional variables.

**Table 3:** This Table looks at the model and setup used by the Harvard study. We run a range of setups and models starting from the main model used in the Harvard study to the main setups and models used in this papers.

| Regression | Dependent | Setup | Counties | Lower Bound | Estimate | Upper Bound |
| --- | --- | --- | --- | --- | --- | --- |
| NBMM | Mortality | Harvard Setup (April 4) | 1785 | 0.0635 | 0.136 | 0.208 |
| NBMM | Mortality | Harvard Setup (April 28) | 3089 | 0.0341 | 0.0897 | 0.145 |
| NBMM | Mortality | Main Setup | 3088 | -0.00101 | 0.0783 | 0.158 |
| NBMM | Mortality | Main Setup + Health | 3029 | -0.00302 | 0.0758 | 0.155 |
| NBMM | Mortality | Main Setup + Transport | 3088 | -0.00853 | 0.0732 | 0.155 |
| NBMM | Mortality | Main Setup + Health + Transport | 3029 | -0.00812 | 0.0731 | 0.154 |
| Linear | Mortality | Harvard Setup (April 4) | 1785 | -29.2 | -14 | 1.07 |
| Linear | Mortality | Harvard Setup (April 28) | 3089 | -2.5 | 15.9 | 34.3 |
| Linear | Mortality per thousand | Main Setup | 3087 | -0.00985 | 0.00424 | 0.0183 |
| Linear | Mortality per thousand | Main Setup + Health + Transport | 3028 | -0.0108 | 0.00227 | 0.0154 |
| Weighted Linear | Mortality | Harvard Setup (April 4) | 1785 | -41.3 | 20.4 | 82.2 |
| Weighted Linear | Mortality | Harvard Setup (April 28) | 3089 | -372 | -147 | 78.8 |
| Weighted Linear | Mortality per thousand | Main Setup | 3087 | -0.0111 | 0.00947 | 0.0301 |
| Weighted Linear | Mortality per thousand | Main Setup + Health + Transport | 3028 | -0.00419 | 0.00875 | 0.0217 |

The remaining set of rows show the estimates from linear regression models using both total deaths as a dependent variable (for the Wu et al. (2020) versions) and deaths per 1,000 estimates for our main setup. In the first four rows, we do not weight counties by their populations; the next four rows do. We start with the same set of counties and covariates as in the April 4 version of Wu et al. (2020). When we estimate this, pollution is negatively correlated with death rates; it is positively correlated with the April 24 version. When we add our additional control variables and health and transport variables, as well as the additional counties, the correlation is no longer negative, but remains statistically insignificant. Weighting the counties by their population tends to make the correlation more positive for the April 4 version, but more makes the result negative for the April 24 version. In both cases this does not change the statistical insignificance of the results.

Taken together, this analysis suggests that the positive correlation between pollution and deaths documented in the April 4 version of Wu et al. (2020) may be driven by the omission of variables that are correlated with both pollution and deaths, not a direct link between pollution and COVID- 19 deaths. The point estimates of the April 24 setup using NBMM are close to those found in our setup, but significance does not seem to hold in our setup. These contrasting results suggest that the correlation between pollution and COVID-19 deaths is not clear and that one should be even more cautious interpreting these correlations as causal relationships.

## Data Availability

All data and code are available.

## Appendix

**Figure 8:**
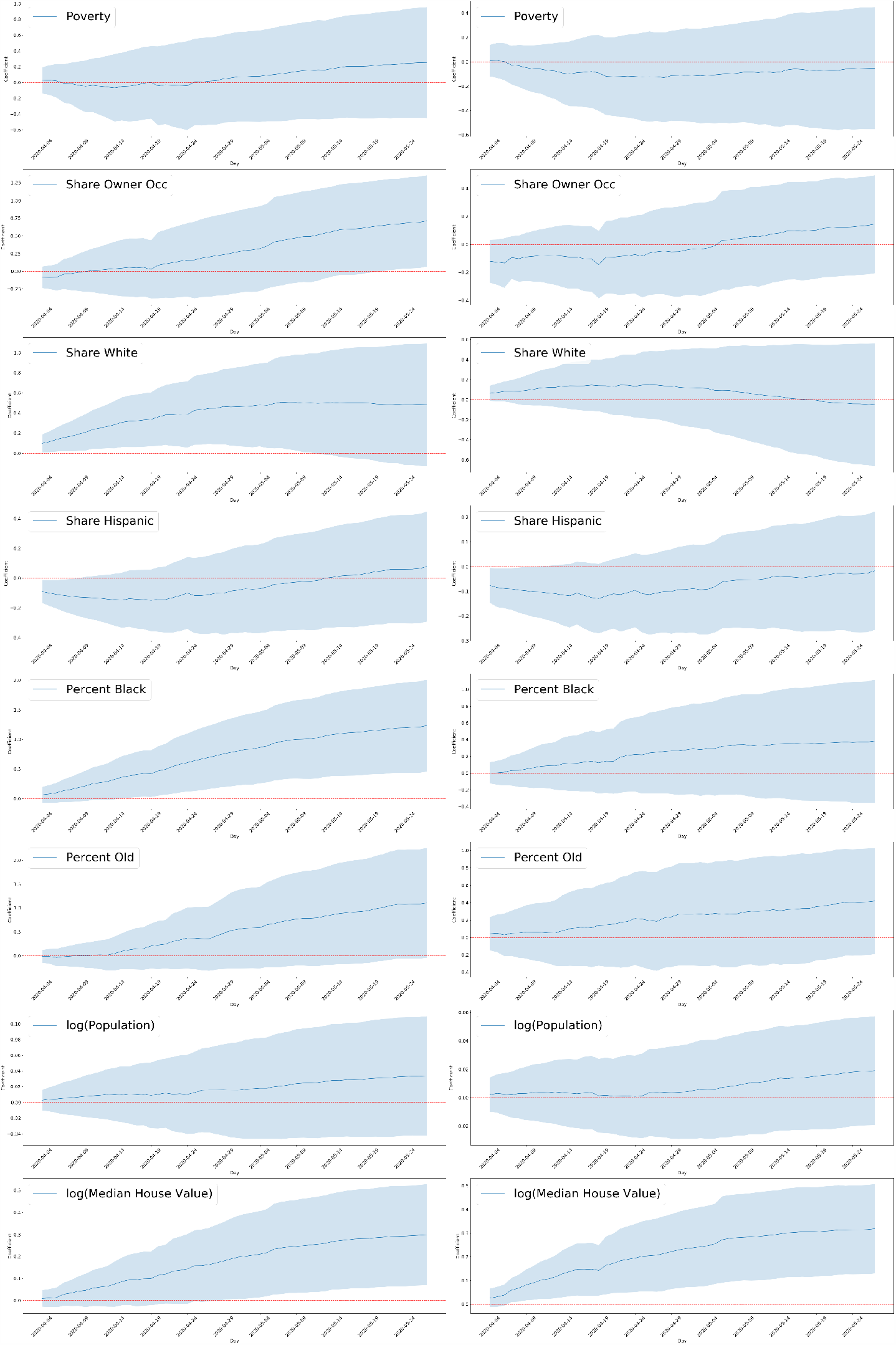
This is a replication of Figure 3 with the additional 5 NY counties.

**Figure 9:**
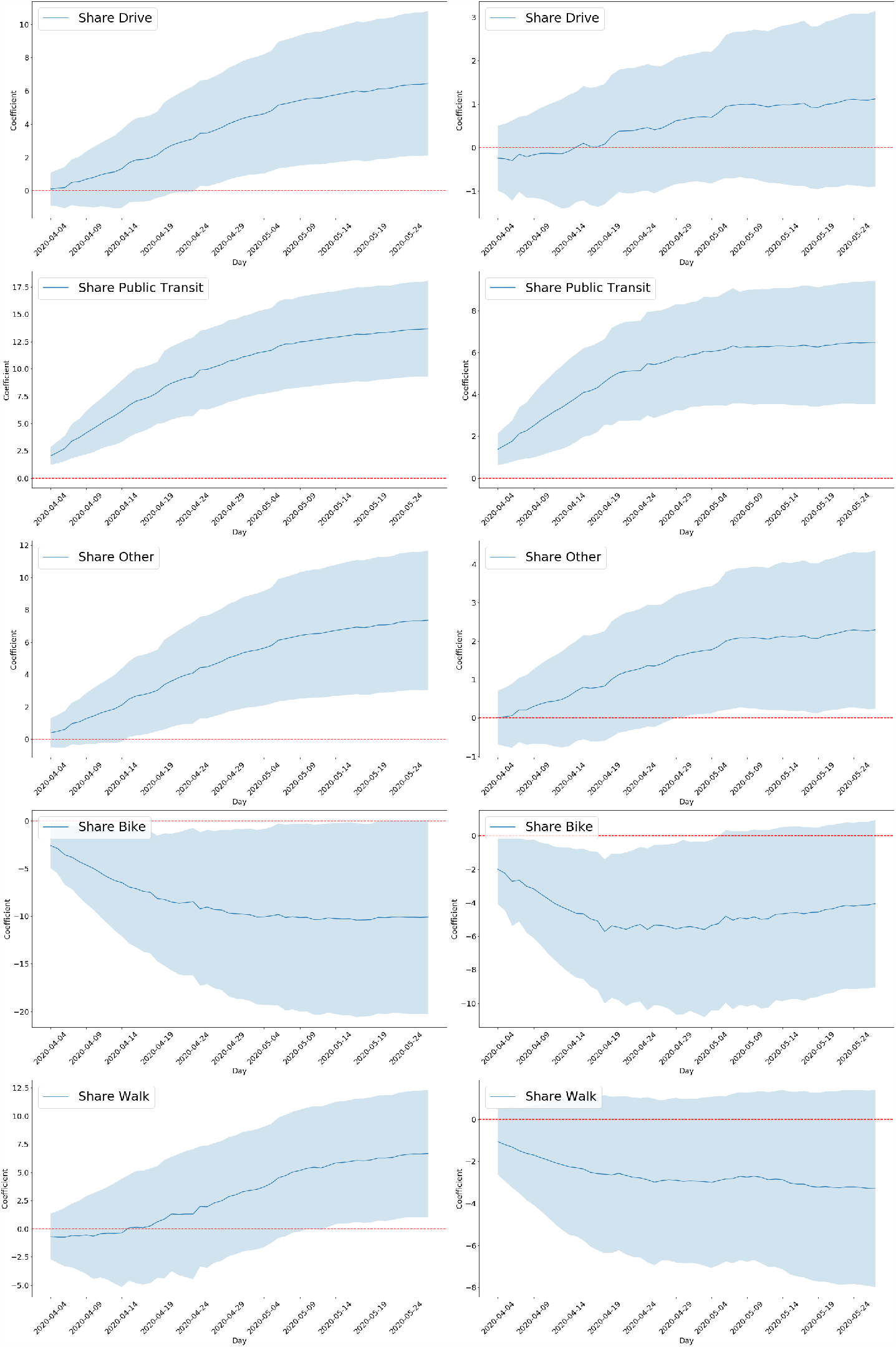
This is a replication of Figure 4 with the additional 5 NY counties.

**Figure 10:**
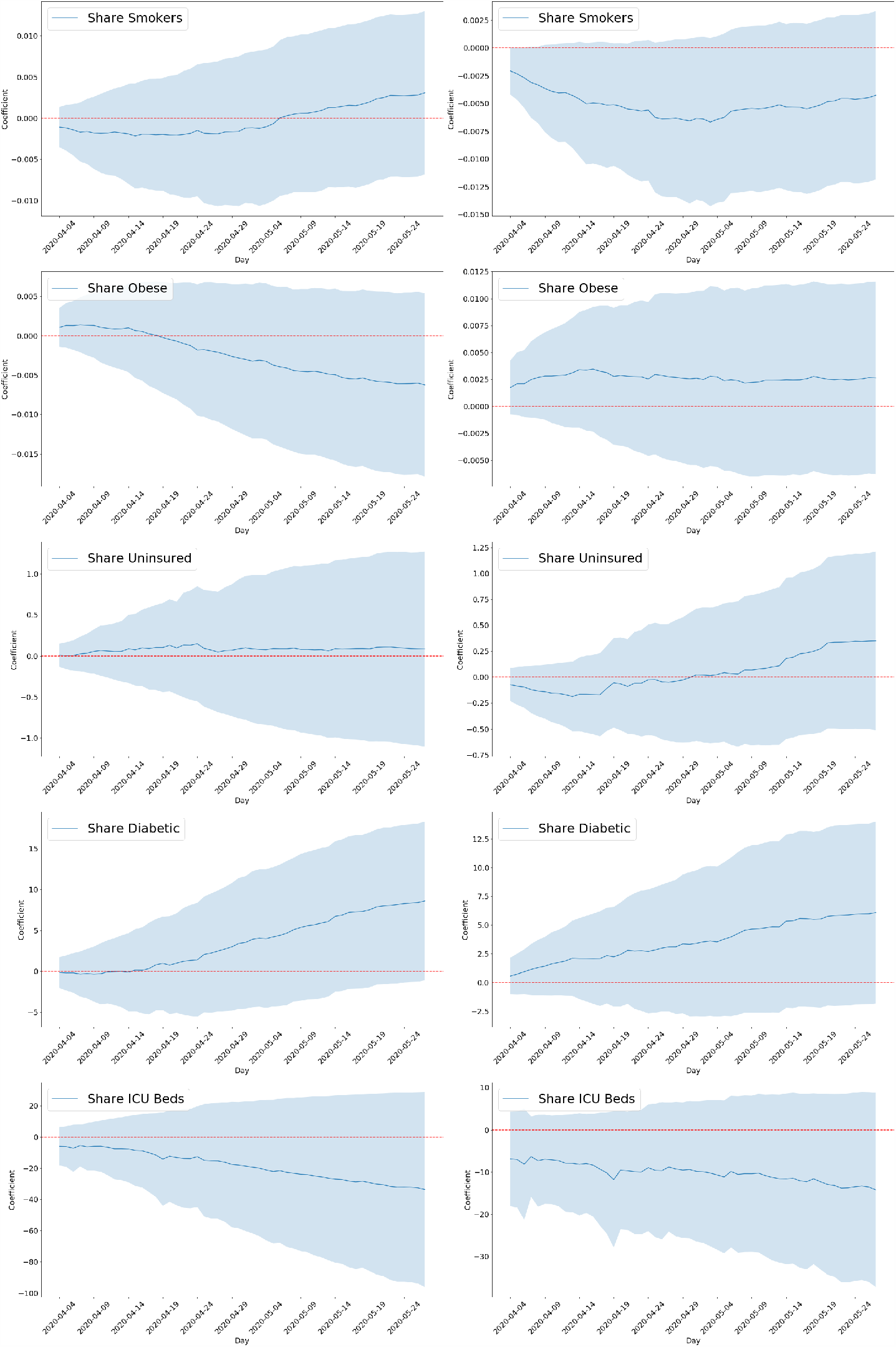
This is a replication of Figure 5 with the additional 5 NY counties.

**Figure 11:**
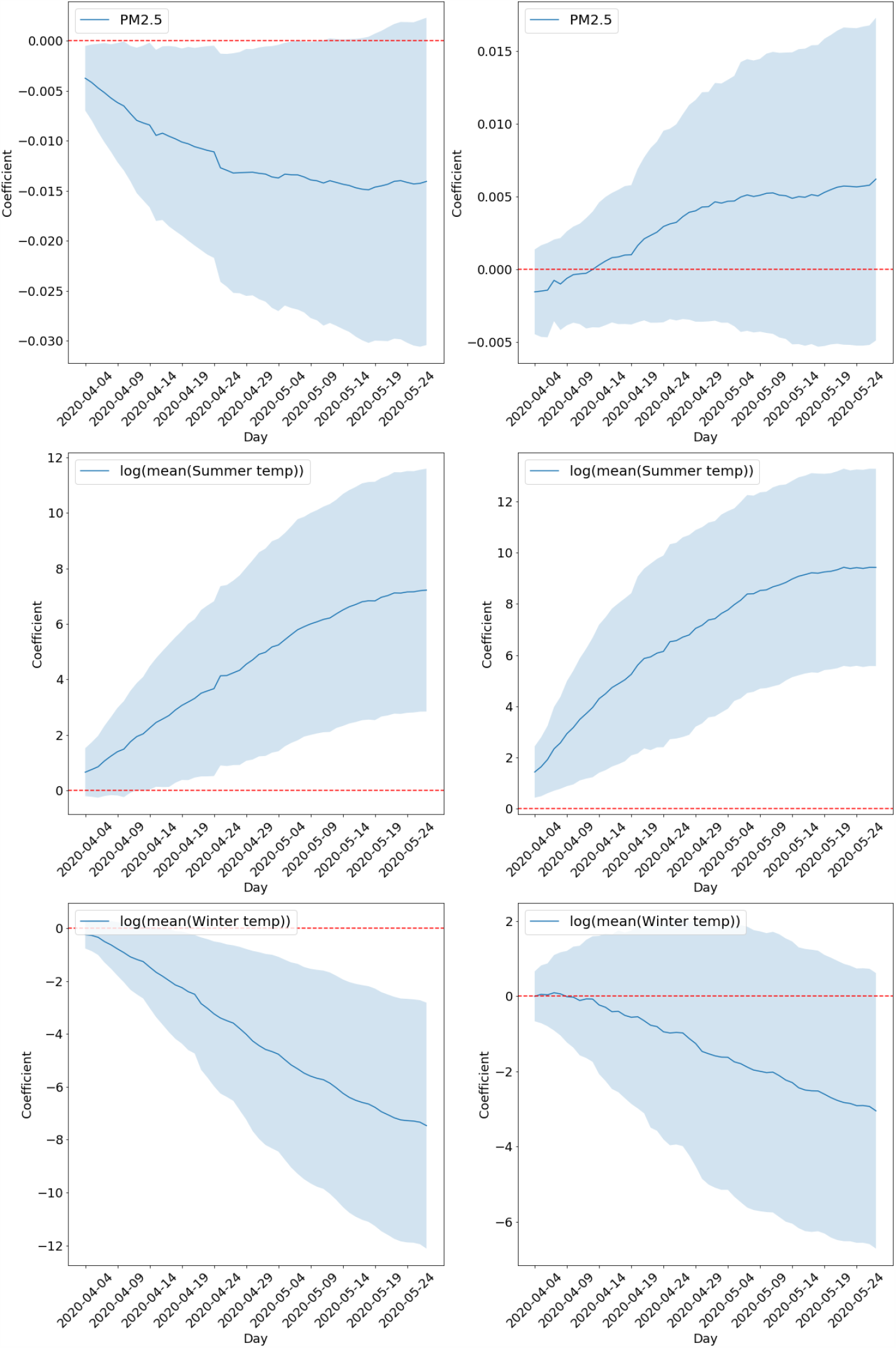
This is a replication of Figure 6 with the additional 5 NY counties.

**Figure 12:**
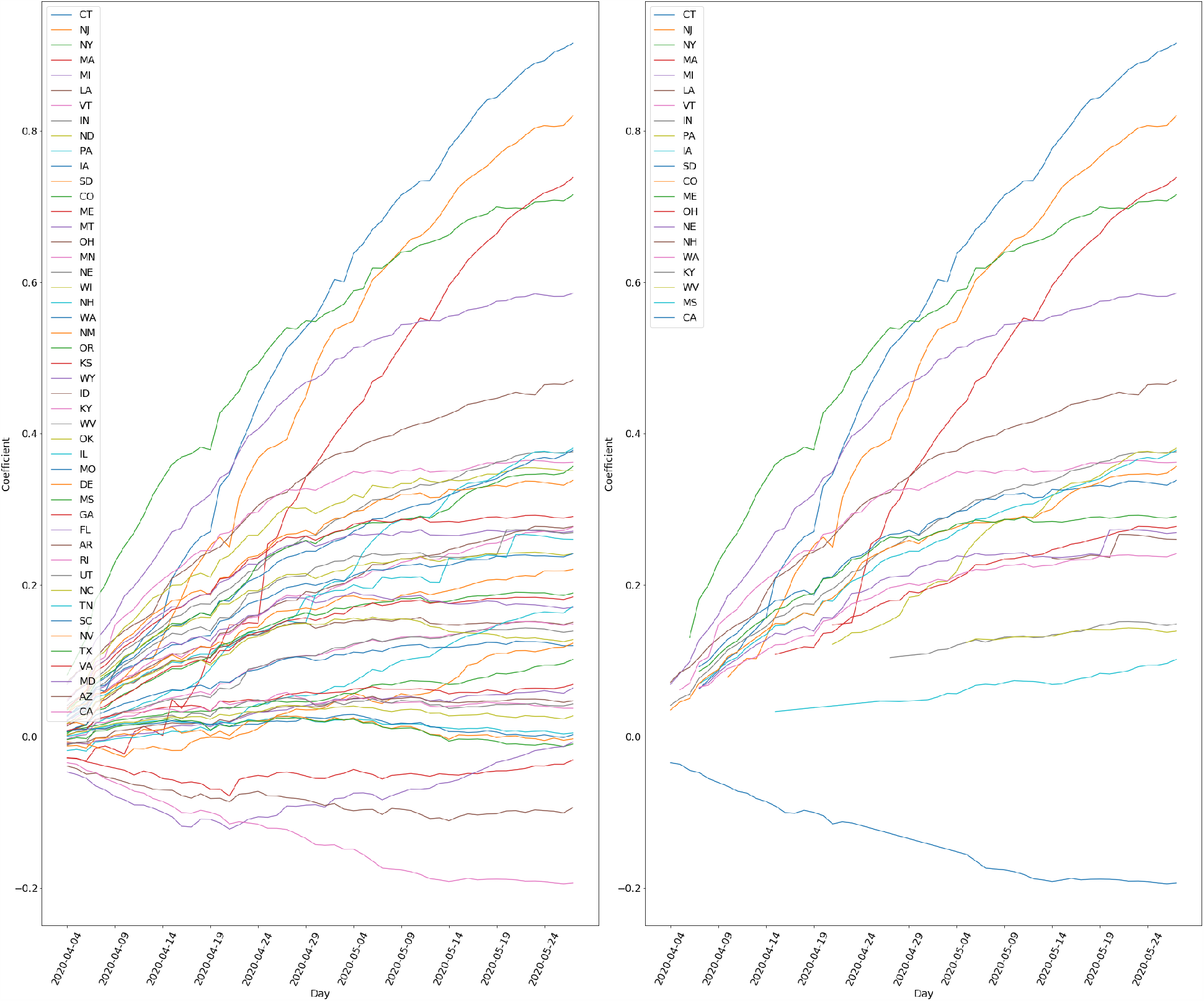
The state Fixed Effect coefficients over time is visualized in this Figure. The subfigure to the left is the visualization for all states, the sugfigure to the right is the visualization of all states that have a coefficient that is significant to the 95% confidence. The state that is dropped in the regression is AL.

**Figure 13:**
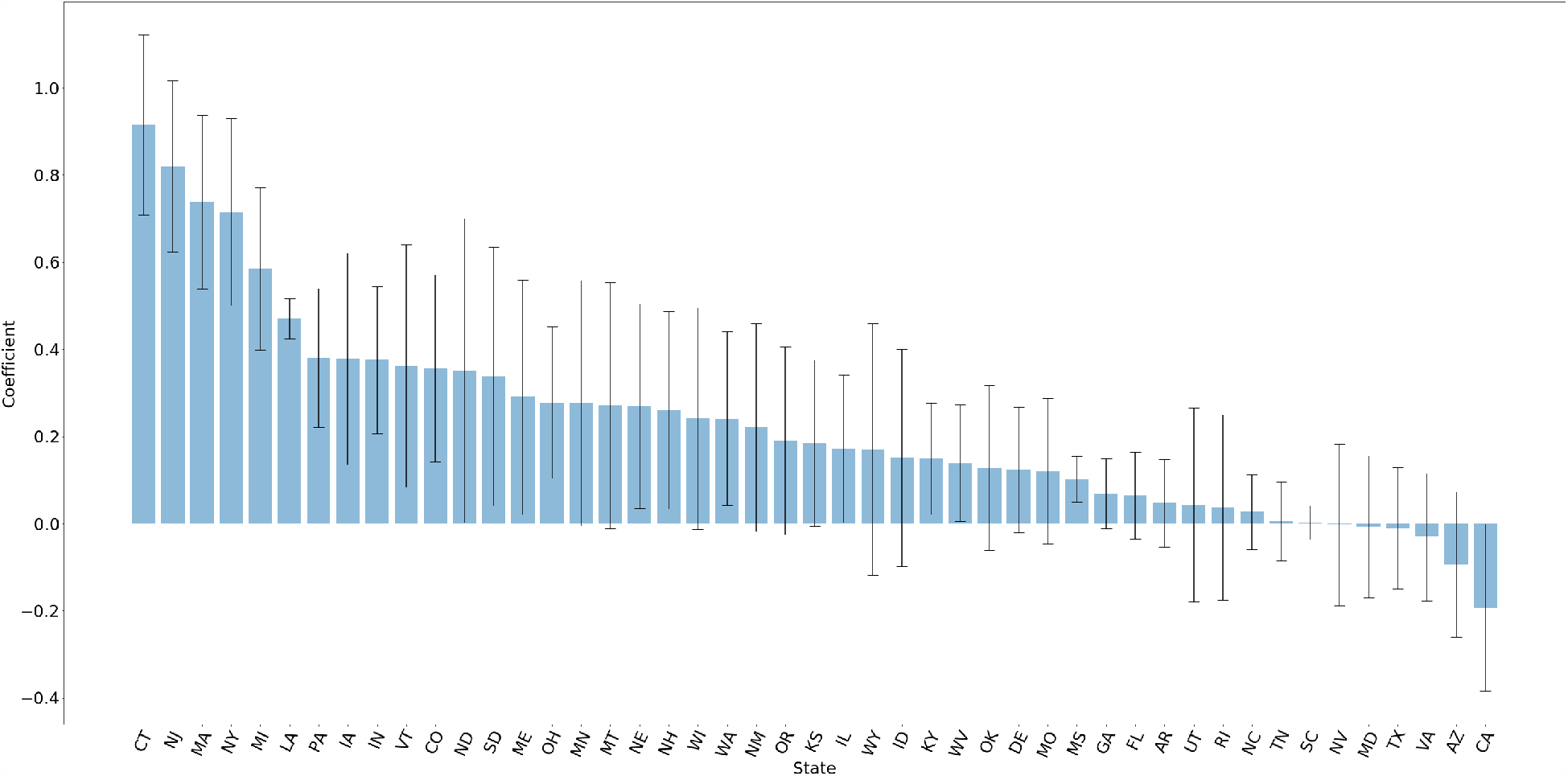
Bar graph of state fixed effects with 95 % CI.

## Footnotes

1 Specifically, our data on mode choice give the number of people using each mode (driving, public transit, walking, biking, and telecommuting) for commuting purposes. We take this number and divide by our population variable to capture the share of the population that is using this commute mode. We de_ne \other” as the di_erence.

2 We are grateful to Wu et al. (2020) for collecting much of the county data we use. We thank the authors for providing their code and data aggregation process online

3 We note that deaths are reported across all five counties rather than individually. Therefore, in the analysis that includes New York City we take a weighted average of the variables included in the model, weighted by the population of each county, and just include one additional observation

4 These results are consistent with recent work by Harris (2020) which correlates drops in subway ridership in New York City with coronavirus doubling rates

5 We can reject, statistically, that each of the coefficients associated with each of the other commute modes equal the coefficient associated with public transit use

6 See, for example: BBC, National Geographic, New York Times, The Guardian

7 We use counties and boroughs interchangeably

8 We are forced to drop 62 counties because of a lack of data on diabetic, ICU beds, and/or older population. Additionally, due to the fact that the 5 boroughs in New York are aggregated into NYC, we drop the 4 boroughs and drop NYC for our main analysis

9 The variables that are in our model as well as the April 4 version of their analysis are: *PM*_2.5_, mean summer and winter temperatures, poverty share, share of owner occupied homes, share of residents that are white and African American, population, share of elderly, mean home value, household income, smoking rate, and education. Their April 24 version adds the obesity share. Our models include the following variables not in their analysis: the commute mode variables, share uninsured, diabetes share, ICU beds per capita, and state indicator variables. Their analysis includes the following variables that we do not: summer and winter average humidity levels, number of hospital beds. Their April 4 version also includes total tests, and BMI, while their April 24 version includes days since the stay-at-home order was announced, days since the first case, share of the population between 45 and 64, and the share of the population between 15 and 44. We take the natural log of income, mean home value, and the summer and winter temperatures, whereas they keep the level. Finally, we include a set of dummy variables for population density, while their April 4 version includes the continuous version of population density

10 This was the most recent analysis on their GitHub repository as of this writing

11 The baseline setup is our socio-economic plus climate and pollution variables.

